# Genitourinary Brucellosis in an Endemic Region: Clinical Spectrum, Diagnostic Pitfalls and Treatment Outcomes of a 13-Year Retrospective Cohort

**DOI:** 10.64898/2026.09.22.26363646

**Authors:** Lütfü Savaş, Abdullah Umut Pekok, Berfin Sude Pekok, Zeynep Sude Eser

## Abstract

**Aim:** Brucellosis remains endemic throughout the Mediterranean basin, the Middle East and Central Asia. Genitourinary involvement is an uncommon but clinically consequential focal complication that closely mimics urological emergencies and may precipitate unnecessary surgery. Contemporary data describing its frequency, laboratory signature and treatment outcomes in endemic settings remain limited.

**Material-Methods:** We retrospectively screened all adults (≥18 years) with confirmed brucellosis managed at a tertiary infectious diseases unit between January 2008 and December 2020. Genitourinary involvement was defined as epididymo-orchitis, orchitis, epididymitis and/or brucellar pyelonephritis. Clinical, laboratory, ultrasonographic, therapeutic and outcome variables were abstracted using a standardised form. Proportions are reported with Wilson score 95% confidence intervals (CI). Reporting followed the STROBE statement.

**Result:** Genitourinary involvement was identified in 18 of 125 patients with brucellosis (14.4%; 95% CI 9.3–21.6); 16 (88.9%) were men and the mean age was 35.2 years (range 22–62). Scrotal disease dominated the male presentation: epididymo-orchitis in 10 of 16 men (62.5%; 95% CI 38.6–81.5), orchitis in 4 (25.0%) and epididymitis in 2 (12.5%). Both women presented with pyelonephritis, one of whom developed a tubo-ovarian abscess requiring drainage. Constitutional features were near-universal (fatigue 100%, night sweats 88.9%, fever 77.8%). Leukocyte counts remained within the reference interval in every patient (mean 5,680/mm3), whereas erythrocyte sedimentation rate (mean 32 mm/h) and C-reactive protein (mean 24.2 mg/L) were only mildly to moderately raised. The Coombs anti-Brucella test was positive in 18 of 18 patients (100%) and the standard agglutination test in 17 of 18 (94.4%), whereas blood culture yielded Brucella melitensis in only 1 of 18 (5.6%). Concomitant extragenitourinary focal disease was present in 8 patients. All patients received doxycycline plus rifampicin; none required modification of the regimen or orchiectomy, and no treatment failure occurred (0 of 18; 95% CI 0–17.6). Median follow-up was 12 weeks (range 6–52) and no relapse was documented within this interval.

**Conclusions:** In endemic regions, genitourinary brucellosis should be suspected in any patient with acute scrotal or flank symptoms accompanied by constitutional complaints, even when the leukocyte count is normal and acute-phase reactants are only marginally elevated. Coombs anti-Brucella serology rather than blood culture is the diagnostic cornerstone, and prompt doxycycline–rifampicin therapy averts surgical intervention.

## INTRODUCTION

Brucellosis is the most widespread bacterial zoonosis worldwide and remains hyperendemic in the Mediterranean basin, the Middle East, Central Asia and parts of Latin America [1, 2]. Transmission occurs through the consumption of unpasteurised dairy products or through occupational contact with infected livestock. Because Brucella species are facultative intracellular pathogens that disseminate haematogenously and persist within the reticuloendothelial system, human brucellosis behaves as a protean systemic illness in which almost any organ may become a focus of infection [3].

Focal complications develop in roughly one third of patients and largely determine morbidity, duration of therapy and the risk of relapse [4]. Osteoarticular disease is by far the most frequent focus, whereas genitourinary involvement has been reported in 2–20% of cases; this wide range reflects heterogeneous case definitions, differing intensity of urological assessment and variable access to scrotal ultrasonography rather than true geographical variation [5–7]. Epididymo-orchitis accounts for the great majority of genitourinary cases in men, while renal parenchymal disease and gynaecological involvement are described almost exclusively in case reports [7, 8].

Genitourinary brucellosis poses a distinctive diagnostic problem. Acute scrotal pain and swelling are indistinguishable from pyogenic epididymo-orchitis, genitourinary tuberculosis or a testicular neoplasm, and the resulting diagnostic delay has repeatedly been linked to unnecessary scrotal exploration and orchiectomy [9, 10]. The difficulty is compounded by a counter-intuitive laboratory profile: leukocytosis is characteristically absent and acute-phase reactants are only modestly raised, so that clinicians who rely on conventional markers of bacterial infection may be falsely reassured [11].

Despite these clinically important pitfalls, the literature is dominated by isolated case reports and small series, and integrated descriptions that link clinical presentation, serological performance, imaging findings and treatment outcome within a single endemic cohort are scarce. We therefore analysed a 13-year consecutive series from a tertiary referral centre in Türkiye with three objectives: (i) to determine the proportion of brucellosis cases with genitourinary involvement and to characterise its clinical spectrum in both sexes; (ii) to describe the laboratory and ultrasonographic signature of the condition, with particular attention to the relative yield of serological and culture-based diagnosis; and (iii) to report the outcome of standard combination antimicrobial therapy, including the need for surgical intervention.

## MATERIALS AND METHODS

### Study design and setting

This was a single-centre, retrospective observational cohort study conducted at the Department of Infectious Diseases and Clinical Microbiology of a tertiary referral hospital serving a brucellosis-endemic catchment area. All patients diagnosed with brucellosis between 1 January 2008 and 31 December 2020 were screened. The study is reported in accordance with the STROBE statement for observational research [12].

### Participants and case definitions

Consecutive adults aged ≥18 years with a confirmed diagnosis of brucellosis were eligible. Brucellosis was diagnosed on the basis of a compatible clinical syndrome together with at least one of the following: a standard tube agglutination test (SAT) titre ≥1:160; a positive Coombs anti-Brucella test; or isolation of Brucella spp. from blood or another sterile site [13]. Patients younger than 18 years, those in whom an alternative aetiology of genitourinary symptoms was established (isolation of a conventional uropathogen, microbiologically or histologically proven tuberculosis, or malignancy) and those with incomplete clinical records were excluded.

Genitourinary involvement was defined as the presence of epididymo-orchitis, orchitis, epididymitis and/or pyelonephritis attributable to brucellosis. Scrotal syndromes were adjudicated as mutually exclusive categories on the basis of combined clinical and ultrasonographic assessment: scrotal pain, swelling and tenderness with sonographic changes confined to the epididymis (epididymitis), confined to the testis (orchitis), or involving both structures (epididymo-orchitis). Pyelonephritis was defined by fever, flank pain and costovertebral angle tenderness together with compatible urinalysis and/or imaging findings in the absence of another uropathogen [7].

Disease duration at presentation was classified as acute (≤8 weeks), subacute (>8–52 weeks) or chronic (>52 weeks) from the onset of symptoms [14]. Acute kidney injury was defined according to the Acute Kidney Injury Network criteria applicable throughout the study period— an increase in serum creatinine of ≥0.3 mg/dL within 48 hours, a ≥50% rise from baseline, or urine output <0.5 mL/kg/h for more than 6 hours — thresholds that are concordant with stage 1 of the later KDIGO classification [15, 16].

Treatment failure was defined a priori as persistence or worsening of clinical findings, persistent elevation of the erythrocyte sedimentation rate (ESR) and C-reactive protein (CRP), new abscess formation, or deterioration of ultrasonographic findings at the end of therapy. Relapse was defined as the recurrence of compatible symptoms with serological or microbiological confirmation within 12 months of completing treatment [17].

### Microbiological and serological methods

Blood cultures were processed with an automated system (BACT/ALERT 3D, bioMérieux, Marcy-l’Étoile, France). Other clinical specimens were inoculated onto standard media and isolates were identified by conventional biochemical methods. Serological evaluation comprised the Rose Bengal plate test, SAT and the Coombs anti-Brucella test; a SAT titre ≥1:160 was regarded as diagnostic in a clinically compatible patient.

### Treatment and follow-up

All patients received doxycycline 100 mg twice daily combined with rifampicin 600–900 mg daily. Treatment duration was individualised according to the site of involvement and clinical response, in line with contemporaneous recommendations for focal brucellosis [18]; the median duration of therapy was seven weeks (range 6-8). Patients were reviewed clinically and with repeat acute-phase reactants for a minimum of 6 weeks after completion of therapy.

### Data collection

Demographic, clinical, laboratory, radiological, therapeutic and outcome variables were abstracted from electronic and paper records onto a standardised case report form by two investigators independently; discrepancies were resolved by consensus. No missing values were present for the variables included in the analysis.

### Statistical analysis

The analysis was descriptive. The distribution of continuous variables was assessed with the Shapiro–Wilk test; normally distributed variables are presented as mean ± standard deviation and non-normally distributed variables as median (range). Categorical variables are presented as counts and percentages with Wilson score 95% confidence intervals, which retain nominal coverage with small denominators. Denominators are stated explicitly for every proportion.

Given that only two women were included, no formal statistical comparison between sexes was undertaken and sex-stratified findings are presented descriptively only. Analyses were performed with IBM SPSS Statistics v26.

### Ethics

The study was approved by the Istinye University Clinical Research Ethics Committee with document number (2017-KAEK-120)/3/2022.G.30 and decision number 3/2022.K-12 (approval no. 2020/145, date: June 15, 2020) and was conducted in accordance with the Helsinki Declaration. Due to the retrospective design and the use of anonymized data, the requirement for individual informed consent was waived by the committee.

## RESULTS

### Cohort and frequency of genitourinary involvement

During the 13-year study period, 125 adults were diagnosed with brucellosis. Genitourinary involvement was identified in 18 of them, corresponding to a frequency of 14.4% (95% CI 9.3–21.6); all 18 eligible patients were included in the final analysis and none were excluded (Figure 1). Sixteen patients were men (88.9%; 95% CI 67.2–96.9) and two were women (11.1%). The mean age was 35.2 years (range 22–62). The duration of the illness at the time of presentation was classified as chronic in all patients. Ten patients (55.6%) had a documented history of consuming unpasteurized dairy products or occupational livestock contact. Baseline characteristics are summarised in Table 1.

**Table 1.** Baseline characteristics, presenting symptoms and clinical findings of patients with genitourinary brucellosis (n = 18).

| Characteristic | n | % (95% CI) |
| --- | --- | --- |
| Male sex | 16 | 88.9 (67.2–96.9) |
| Female sex | 2 | 11.1 (3.1–32.8) |
| Age, years — mean (range) | — | 35.2 (22–62) |
| **Constitutional symptoms** |  |  |
| Fatigue | 18 | 100 (82.4–100) |
| Night sweats | 16 | 88.9 (67.2–96.9) |
| Fever | 14 | 77.8 (54.8–91.0) |
| Anorexia | 12 | 66.7 (43.7–83.7) |
| Back pain | 12 | 66.7 (43.7–83.7) |
| Arthralgia | 11 | 61.1 (38.6–79.7) |
| Weight loss | 10 | 55.6 (33.7–75.4) |
| **Genitourinary symptoms and signs** |  |  |
| Testicular tenderness (men, n = 16) | 16 | 100 (80.6–100) |
| Scrotal swelling (men, n = 16) | 15 | 93.8 (71.7–98.9) |
| Scrotal hyperaemia (men, n = 16) | 14 | 87.5 (64.0–96.5) |
| Dysuria (all, n = 18) | 4 | 22.2 (9.0–45.2) |
| Haematuria (all, n = 18) | 1 | 5.6 (1.0–25.8) |
| Urethral discharge (men, n = 16) | 1 | 6.3 (1.1–28.3) |
| **Comorbidity** |  |  |
| Diabetes mellitus | 1 | 5.6 (1.0–25.8) |
| Hypertension | 1 | 5.6 (1.0–25.8) |
| Urolithiasis | 1 | 5.6 (1.0–25.8) |
| Benign prostatic hyperplasia | 1 | 5.6 (1.0–25.8) |
| Immunosuppression | 0 | 0 (0–17.6) |

*CI, confidence interval. Percentages are calculated on the denominator stated in parentheses for each variable; Wilson score confidence intervals are shown.*

**Figure 1.**
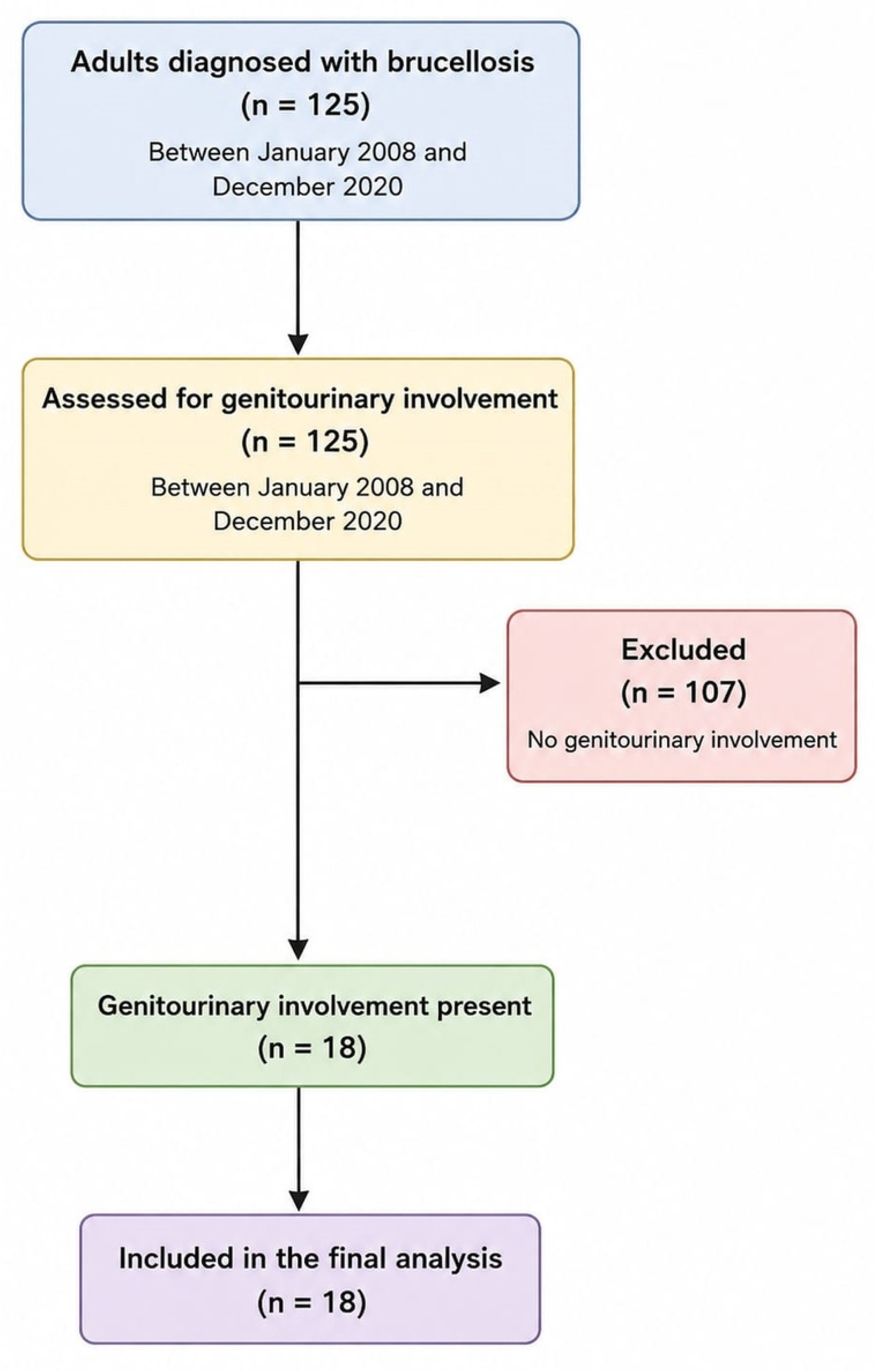
Flow of patients through the study. Of 125 adults diagnosed with brucellosis between January 2008 and December 2020, 18 had genitourinary involvement and all were included in the final analysis.

### Clinical presentation

Constitutional symptoms dominated the presentation and were present in nearly all patients: fatigue in 18 (100%), night sweats in 16 (88.9%), fever in 14 (77.8%), anorexia in 12 (66.7%), back pain in 12 (66.7%), arthralgia in 11 (61.1%) and weight loss in 10 (55.6%) (Table 1).

Among the 16 male patients, local scrotal findings were uniformly present: testicular tenderness in 16 (100%), scrotal swelling in 15 (93.8%) and scrotal hyperaemia in 14 (87.5%). Both female patients presented with the clinical picture of acute pyelonephritis and both reported dysuria; one also had macroscopic haematuria. Dysuria was additionally reported by two men, and one man had urethral discharge.

Comorbidities were infrequent and comprised diabetes mellitus, urolithiasis, benign prostatic hyperplasia and hypertension in one patient each (5.6% each). No patient had an immunosuppressive condition or was receiving immunosuppressive therapy, and neither of the two women was pregnant.

### Laboratory and serological findings

The leukocyte count remained within the reference interval in every patient (mean 5,680/mm3), and neither anaemia nor thrombocytopenia was prominent (mean haemoglobin 12.7 g/dL; mean platelet count 255,000/mm3). By contrast, acute-phase reactants were mildly to moderately elevated, with a mean ESR of 32 mm/h and a mean CRP of 24.2 mg/L (Table 2).

**Table 2.** Laboratory findings at presentation (n = 18).

| Parameter | Mean ± SD | Median (range) | Reference interval |
| --- | --- | --- | --- |
| Leukocytes, /mm<sup>3</sup> | 5,680 ± 1,280 | 6,570 (5,500-10,500) | 4,000–11,000 |
| Haemoglobin, g/dL | 12.7 ± 1.2 | 12 (11-14) | 12–18 |
| Platelets, /mm<sup>3</sup> | 255,000 ± 25,000 | 185,000 (155,000-325,000) | 150,000–450,000 |
| ESR, mm/h | 32 ± 8.4 | 31 (range 18-52) | ≤20 |
| CRP, mg/L | 24.2 ± 5.8 | 28 (range 22-64) | 0–5 |
| Serum creatinine, mg/dL | 1.1 ± 0.2 | 1.0 (range 0.9-1.4) | 0.6–1.2 |
| Patients with leukocytosis, n (%) | 0 (0%) | — | — |
| Patients with elevated ESR, n (%) | 18 (100%) | — | — |
| Patients with elevated CRP, n (%) | 18 (100%) | — | — |

*CRP, C-reactive protein; ESR, erythrocyte sedimentation rate; SD, standard deviation. Cells highlighted in the working copy must be completed from the source dataset before submission; Q1 journals will not accept measures of central tendency reported without a measure of dispersion.*

Serological testing established the diagnosis in all cases. The Coombs anti-Brucella test was positive in 18 of 18 patients (100%; 95% CI 82.4–100), SAT at a titre ≥1:160 in 17 of 18 (94.4%; 95% CI 74.2–99.0) and the Rose Bengal plate test in 16 of 18 (88.9%; 95% CI 67.2– 96.9). Microbiological confirmation was much less sensitive: Blood culture was positive in only 1 of 18 patients (5.6%; 95% CI 1.0–25.8) and Brucella melitensis was detected; in contrast, urine and genital discharge cultures were negative in all patients.

### Ultrasonographic findings

Scrotal ultrasonography demonstrated inflammatory change in 15 of 16 men (93.8%; 95% CI 71.7–98.9) and testicular involvement in 12 (75.0%; 95% CI 50.5–89.8). Abdominal ultrasonography showed hepatomegaly in 4 patients (22.2%) and splenomegaly in 4 (22.2%). No testicular or renal abscess was demonstrated in any patient at presentation (Table 3).

**Table 3.**
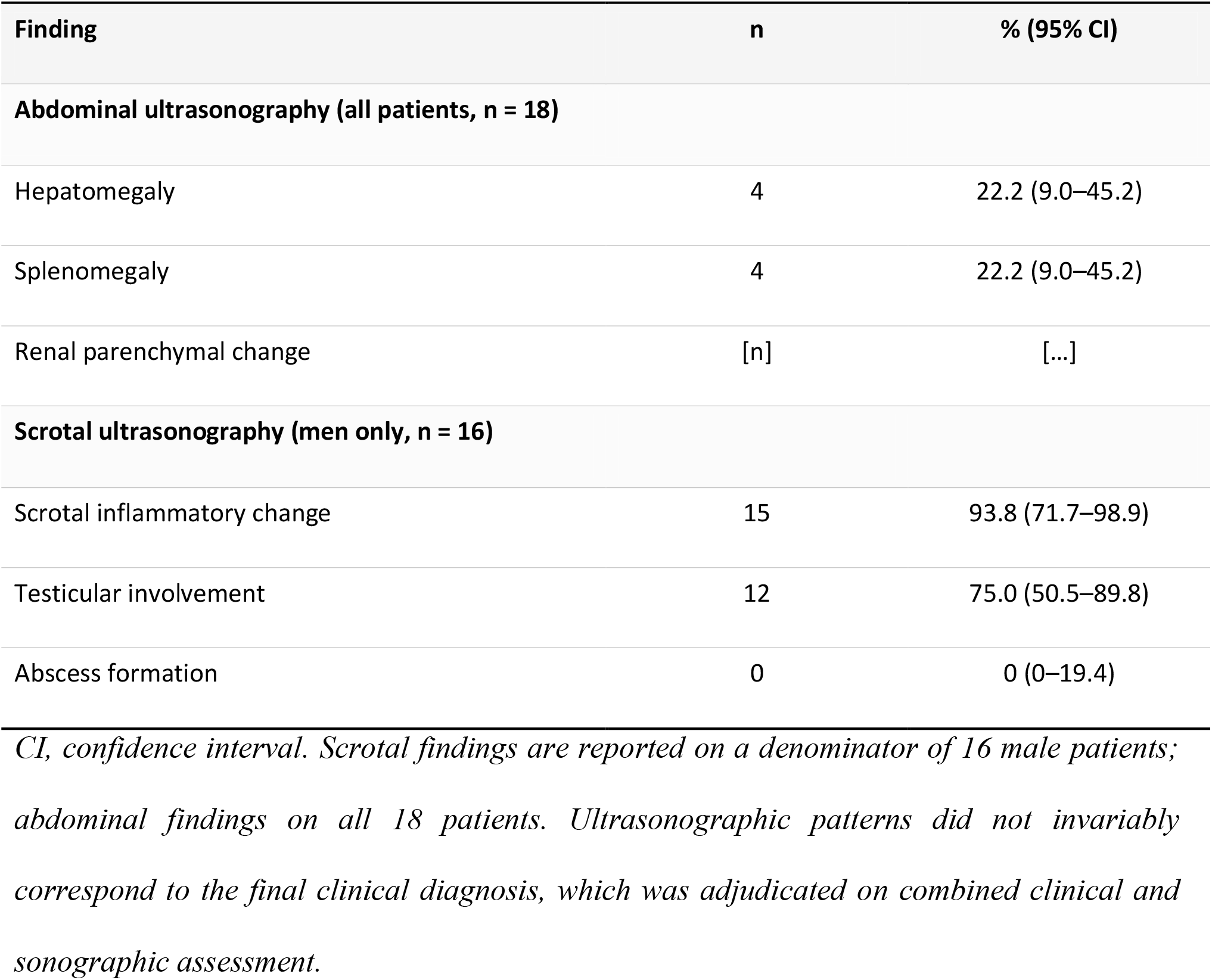
Ultrasonographic findings.

| Finding | n | % (95% CI) |
| --- | --- | --- |
| **Abdominal ultrasonography (all patients, n = 18)** |  |  |
| Hepatomegaly | 4 | 22.2 (9.0–45.2) |
| Splenomegaly | 4 | 22.2 (9.0–45.2) |
| Renal parenchymal change | [n] | [...] |
| **Scrotal ultrasonography (men only, n = 16)** |  |  |
| Scrotal inflammatory change | 15 | 93.8 (71.7–98.9) |
| Testicular involvement | 12 | 75.0 (50.5–89.8) |
| Abscess formation | 0 | 0 (0–19.4) |

*CI, confidence interval. Scrotal findings are reported on a denominator of 16 male patients; abdominal findings on all 18 patients. Ultrasonographic patterns did not invariably correspond to the final clinical diagnosis, which was adjudicated on combined clinical and sonographic assessment.*

### Final diagnoses and concomitant focal involvement

Among the 16 men, the final adjudicated diagnosis was epididymo-orchitis in 10 (62.5%; 95% CI 38.6–81.5), orchitis in 4 (25.0%; 95% CI 10.2–49.5) and epididymitis in 2 (12.5%; 95% CI 3.5–36.0). Both women were diagnosed with brucellar pyelonephritis, representing 2 of 18 patients overall (11.1%) (Table 4).

**Table 4.**
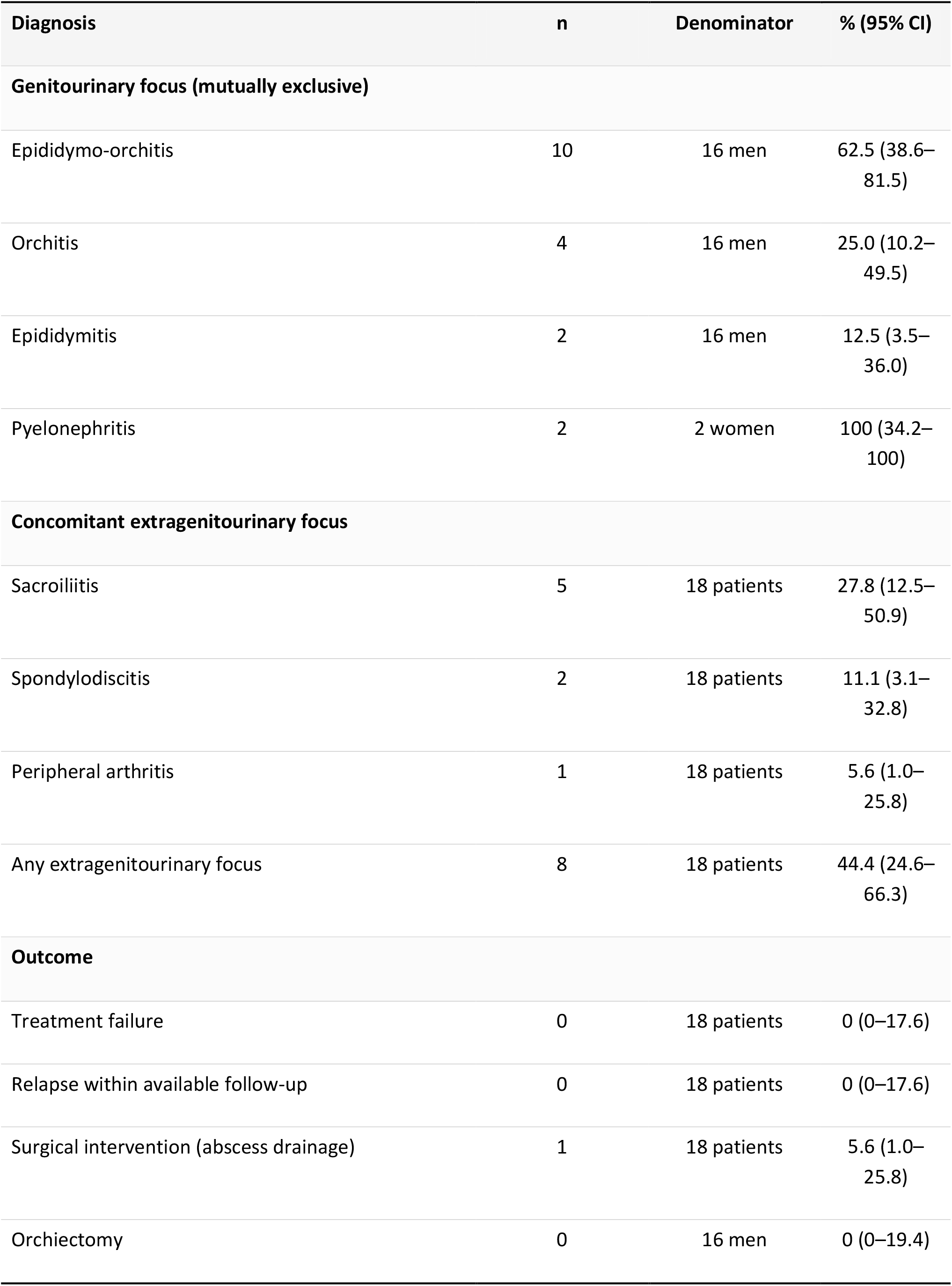
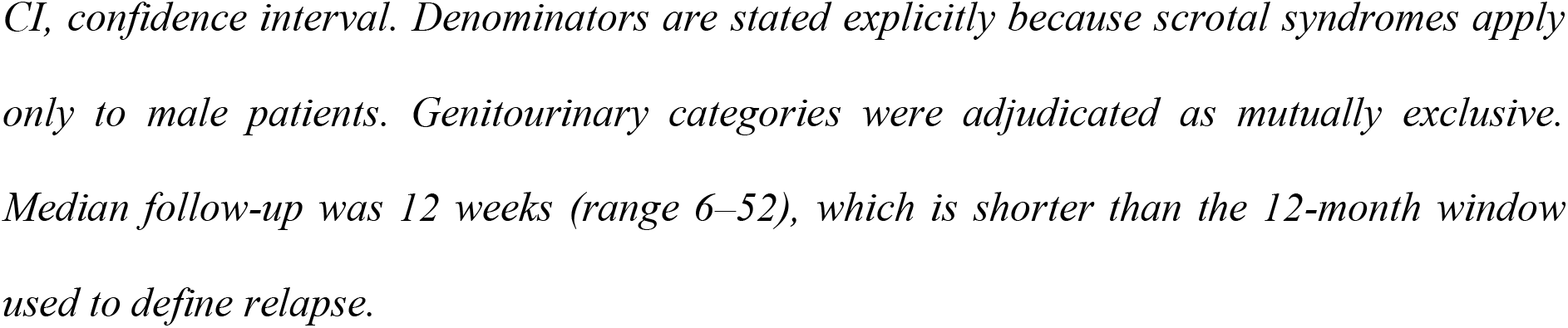
Final adjudicated diagnoses and concomitant focal involvement.

| Diagnosis | n | Denominator | % (95% CI) |
| --- | --- | --- | --- |
| **Genitourinary focus (mutually exclusive)** |  |  |  |
| Epididymo-orchitis | 10 | 16 men | 62.5 (38.6–81.5) |
| Orchitis | 4 | 16 men | 25.0 (10.2–49.5) |
| Epididymitis | 2 | 16 men | 12.5 (3.5–36.0) |
| Pyelonephritis | 2 | 2 women | 100 (34.2–100) |
| **Concomitant extragenitourinary focus** |  |  |  |
| Sacroiliitis | 5 | 18 patients | 27.8 (12.5–50.9) |
| Spondylodiscitis | 2 | 18 patients | 11.1 (3.1–32.8) |
| Peripheral arthritis | 1 | 18 patients | 5.6 (1.0–25.8) |
| Any extragenitourinary focus | 8 | 18 patients | 44.4 (24.6–66.3) |
| **Outcome** |  |  |  |
| Treatment failure | 0 | 18 patients | 0 (0–17.6) |
| Relapse within available follow-up | 0 | 18 patients | 0 (0–17.6) |
| Surgical intervention (abscess drainage) | 1 | 18 patients | 5.6 (1.0–25.8) |
| Orchiectomy | 0 | 16 men | 0 (0–19.4) |

*CI, confidence interval. Denominators are stated explicitly because scrotal syndromes apply only to male patients. Genitourinary categories were adjudicated as mutually exclusive. Median follow-up was 12 weeks (range 6–52), which is shorter than the 12-month window used to define relapse.*

Focal involvement outside the genitourinary tract coexisted in 8 of 18 patients (44.4%): sacroiliitis in 5 (27.8%; 95% CI 12.5–50.9), spondylodiscitis in 2 (11.1%) and peripheral arthritis in 1 (5.6%). No patient had endocarditis or neurobrucellosis.

### Treatment and outcomes

All 18 patients (100%) were treated with the doxycycline–rifampicin combination and none required modification of the initial regimen because of intolerance or inadequate response. No treatment failure was recorded (0 of 18; 95% CI 0–17.6). One woman developed a tubo-ovarian abscess during follow-up and underwent surgical drainage; because she had already demonstrated a clinical and biochemical response to antimicrobial therapy, this event was adjudicated as a complication of the disease rather than as treatment failure. No man underwent scrotal exploration or orchiectomy.

The median length of hospital stay was 9.5 days (range 2–20). No patient developed acute kidney injury during treatment or follow-up. Median follow-up after completion of therapy was 12 weeks (range 6–52), and no relapse was documented within the available follow-up interval; the implications of this follow-up duration are addressed in the Limitations section.

## DISCUSSION

In this 13-year consecutive cohort from an endemic region, genitourinary involvement complicated 14.4% of adult brucellosis cases and presented in a strongly sex-dependent manner: scrotal disease, and specifically epididymo-orchitis, accounted for the entire male presentation, whereas both women presented with pyelonephritis, one of whom developed a tubo-ovarian abscess. Three findings have direct clinical implications. First, the systemic inflammatory response was strikingly muted, with a normal leukocyte count in every patient. Second, serology — and in particular the Coombs anti-Brucella test — vastly outperformed culture, which was positive in fewer than one in ten patients. Third, uniform treatment with doxycycline and rifampicin produced clinical resolution in all patients without any surgical intervention on the scrotum.

The observed frequency of 14.4% lies at the upper end of the 2–20% range reported in the literature and exceeds the figure of approximately 10% that is often quoted [5, 6]. Two explanations are plausible. Our institution is a tertiary referral centre to which patients with focal complications are preferentially directed, so ascertainment bias is likely; conversely, scrotal ultrasonography was performed liberally in symptomatic men, which may have increased the detection of mild epididymal involvement that would otherwise have been missed. The distribution of scrotal syndromes we observed is nevertheless closely concordant with previous series. Navarro-Martínez et al. described epididymo-orchitis in 59 patients with Brucella melitensis infection and Memish and Venkatesh reported a comparable predominance from Saudi Arabia [7, 9], while the Turkish multicentre study by Erdem et al [19] and the 1,028-patient series of Buzgan et al [20]. Both identified epididymo-orchitis as the leading genitourinary manifestation with low relapse rates after adequate therapy [19, 20].

The dissociation between evident organ involvement and an unremarkable leukocyte count deserves emphasis because it is a recurrent cause of diagnostic delay. This pattern is biologically coherent. Brucella species are facultative intracellular organisms that survive within macrophages, and their lipopolysaccharide has markedly lower endotoxic activity than that of enteric Gram-negative bacilli, eliciting a granulomatous rather than a pyogenic host response with limited neutrophil recruitment [2, 3]. The practical consequence is that a normal white cell count carries essentially no negative predictive value in this setting, and that a syndromic approach integrating epidemiological exposure, constitutional symptoms and organ-specific findings must take precedence over conventional inflammatory markers — particularly in resource-constrained endemic settings where reliance on such markers may delay effective therapy.

Our serological data reinforce a second practical message. The Coombs anti-Brucella test was positive in every patient, whereas SAT missed one case and blood culture identified the organism in only one. Blocking (incomplete) IgG and IgA antibodies, which are not detected by standard agglutination and which accumulate in longer-standing or focal disease, are captured by the Coombs technique, and prozone phenomena may further reduce SAT sensitivity at low dilutions [13]. Blood culture yield, in turn, is heavily dependent on prior antimicrobial exposure and on disease duration and is characteristically low in focal, non-bacteraemic presentations. Clinicians investigating suspected genitourinary brucellosis should therefore not regard a negative SAT or a negative blood culture as sufficient grounds for exclusion, and Coombs testing should be requested explicitly.

Because brucellar epididymo-orchitis is clinically and sonographically indistinguishable from pyogenic epididymo-orchitis, genitourinary tuberculosis and testicular neoplasia, the differential diagnosis carries direct surgical consequences: relapsing brucellosis simulating a testicular tumour and brucellar testicular abscess presenting as a scrotal mass have both been reported, and unnecessary orchiectomy remains a real hazard [8, 10]. Our ultrasonographic data illustrate the limits of imaging: sonographic patterns did not map consistently onto the final adjudicated diagnosis, and scrotal ultrasonography is best regarded as a means of excluding abscess, torsion and tumour rather than as a means of establishing an aetiology. No patient in our series required scrotal exploration, and we consider this to be the most clinically relevant consequence of early aetiological recognition. Delayed or inadequate treatment, by contrast, may lead to abscess formation, testicular atrophy, segmental infarction, impaired spermatogenesis and infertility [21–24].

Genitourinary brucellosis in women is far less well characterised. Both of our female patients presented with pyelonephritis and one developed a tubo-ovarian abscess requiring drainage — a rare but well-recognised gynaecological complication. Renal involvement in brucellosis spans interstitial nephritis, glomerulonephritis and, rarely, renal brucelloma, and may occur in either sex, sometimes in association with endocarditis [11]; no such association was present in our cohort, and no patient developed acute kidney injury. With only two women, our data cannot support inference about sex-related differences in pathogenesis or outcome and are reported descriptively. They do, however, support the pragmatic recommendation that brucellosis be considered in women from endemic areas who present with pyelonephritis or a pelvic inflammatory syndrome that fails to respond to conventional therapy, especially given the well-documented association of brucellosis in pregnancy with miscarriage, preterm delivery and intrauterine infection [25].

From a therapeutic standpoint, the intracellular localisation of Brucella within the reticuloendothelial system limits antimicrobial penetration and mandates prolonged combination therapy. The uniform response to doxycycline plus rifampicin in our cohort supports the continued use of this regimen as first-line therapy for uncomplicated genitourinary disease, and its oral administration is a considerable practical advantage in outpatient management. This result should nevertheless be interpreted with caution. Rifampicin induces the metabolism of doxycycline and lowers its serum concentrations, and randomised and pooled analyses have suggested that doxycycline combined with an aminoglycoside is associated with fewer relapses than doxycycline–rifampicin, particularly in focal disease [17, 18, 26]. Our observation of no failures should therefore not be read as evidence of equivalence; it reflects a small cohort of predominantly uncomplicated, early-recognised disease followed for a limited period.

### Limitations

Several limitations must be acknowledged. The study was retrospective and conducted at a single tertiary centre, which introduces both referral and ascertainment bias and limits generalisability. The sample was small; because genitourinary involvement is an uncommon manifestation, only 18 patients accrued over 13 years, and the resulting confidence intervals around all proportions are wide. Only two women were included, so no meaningful sex-based comparison was possible and the female findings should be regarded as observational only. Critically, no comparison group of brucellosis patients without genitourinary involvement was assembled; consequently the study cannot identify risk factors for genitourinary dissemination, and this remains the principal analytical gap. The median follow-up of 12 weeks was substantially shorter than the 12-month window used to define relapse, so the absence of documented relapse almost certainly underestimates the true relapse rate and this outcome should be regarded as provisional. Finally, no patient underwent semen analysis or long-term fertility assessment, and no molecular typing or minimum inhibitory concentration testing of the single isolate was performed.

## CONCLUSION

Genitourinary involvement complicated approximately one in seven cases of adult brucellosis in this endemic cohort, presenting as epididymo-orchitis in men and as pyelonephritis in women. A normal leukocyte count and only marginally elevated acute-phase reactants are the rule rather than the exception and must not be allowed to exclude the diagnosis. Coombs anti-Brucella serology, not blood culture, is the diagnostic cornerstone. Where the diagnosis is made early, oral doxycycline–rifampicin therapy resolves the illness without recourse to scrotal surgery. Prospective, multicentre studies incorporating a non-genitourinary comparison group, follow-up extending to at least 12 months, and assessment of fertility outcomes are required to define risk factors for genitourinary dissemination and to establish the optimal regimen.

## Data Availability

The anonymised dataset supporting the findings of this study is available from the corresponding author on reasonable request, subject to institutional and ethical restrictions protecting patient confidentiality.

## DECLARATIONS

Ethics approval and consent to participate. Approved by the Institutional Ethics Committee of Istinye University (2020/145, 15 June 2020). Conducted in accordance with the Declaration of Helsinki. Informed consent was waived owing to the retrospective design. Consent for publication. Not applicable; no individually identifiable data are presented. Availability of data and materials. The anonymised dataset supporting the conclusions of this article is available from the corresponding author on reasonable request. Competing interests. The authors declare that they have no competing interests. research received no specific grant from any funding agency in the public, commercial or not-for-profit sectors.

Authors’ contributions (CRediT). LS: conceptualisation, methodology, investigation, writing − original draft, supervision. AUP: investigation, data curation, formal analysis, writing – review and editing. BSP: investigation, data curation, formal analysis, writing – review and editing. ZSE: investigation, data curation, formal analysis, writing – review and editing. All authors read and approved the final manuscript.

## Acknowledgements

None.

